# Exploring virtual funding committee practices in the allocation of National Institute for Health and Care Research funding: A netnographic study

**DOI:** 10.1101/2023.11.17.23298707

**Authors:** Amanda Jane Blatch-Jones, Cherish Boxall, Emmanuel Asante, Katie Meadmore

## Abstract

**Objectives:** Funding committees, comprising members with a range of knowledge, skills, and experience, are considered integral to the decision-making process of funding organisations for recommending or allocating research funding. However, there is limited research investigating the decision-making processes, the role of members and their social interactions during funding committee meetings conducted both virtually and face-to-face.

**Methods:** Using a mixed-methods design and following netnography principles, the study observed nine National Institute for Health and Care Research programmes funding committee meetings conducted virtually during October 2020 to December 2021; complemented by interviews with committee chairs and members (18 interviews) and NIHR staff (12 interviews); an online survey (50 responses); and documentary analysis. Personal reflections through immersive journals also formed part of the analysis.

**Results:** Three main themes were identified from the observations, interviews, and online survey: ***efficiency of virtual committee meetings*** (importance of preparation, and the role of formality, process, and structure); ***understanding the effect of virtual committee meetings on well-being*** (effects of fatigue and apprehension, and the importance of work life balance); and, ***understanding social interactions and engagement*** (levels of engagement, contribution and inclusivity, awareness of unconscious bias and the value of social networking).

**Conclusions:** Examining the decision-making practices of one funding organisation across several research programmes, across multiple committee meetings over one year has generated new insights around funding committee practices that previous studies have not been able to explore or investigate. Overall, it was observed that fair and transparent funding recommendations and outcomes can be achieved through virtual funding committees. However, whilst virtual funding committees have many benefits and opportunities, such as the potential to increase membership diversity and inclusivity, and be more environmentally sustainable, more evidence is needed to evaluate their effectiveness, with particular focus on issues of fatigue, engagement, and committee cohesion, especially when new committee members join.

## INTRODUCTION

Funding organisations rely on decision-making procedures to support them to make funding recommendations that are effective, fair, and transparent.^(1)^ An integral part of the process involves members with a range of knowledge, skills, and experience (often referred to as funding committees or panels) who convene to evaluate and recommend the allocation of research funding. Several assessments and processes are carried out to support and enable funding committee decision-making. For example, using external peer reviewers to offer an impartial, independent review that informs the funding committee process for funding allocation.^(2, 3)^ Despite the valuable role these committees play to ensure quality, fair and transparent allocation of research funding, there is a lack of empirical evidence on the processes and functions of funding committee practices. For example, Guthrie *et al.*^(4)^ found no studies examining the social processes of funding committees, despite their central role in the funding allocation process. This could be related to the sensitivity and accessibility around the funding allocation processes and procedures of funding organisations (e.g., funding committee discussions and confidentiality of research applications).^(2, 3, 5–7)^ Challenges in gaining access to funding committees to undertake research or through direct observations is also reported in the literature, along with a lack of well-conducted research looking at more than one funding organisation or in more than one particular context (e.g., more than one research grant programme).^(4, 5)^

### Virtual interactions

In 2020, the unprecedented global COVID-19 pandemic challenged the conduct of face-to-face funding committee meetings. This resulted in rapid changes to how funding organisations continued the assessment of research applications, whilst maintaining quality, transparency and fairness of their research funding practices. Whilst most of the evidence focuses on committee members scoring and how videoconferencing may influence final decision-making scores of funding panels, ^(2, 4, 8–10)^ there is limited literature on the use of virtual online platforms as an alternative to face-to-face meetings. For example, Pier *et al.* examined the degree of scoring variability across different panels and whether there were differences between videoconferencing to in-person peer review of research proposals, as they did not have access to actual National Institutes of Health (NIH) study designs.^(9)^ They found minimal variation on the final scores between videoconference and in-person meetings, which also supported Gallo *et al.’s* earlier findings that most review outcomes are not affected by the review setting.^(11)^

Attempts to understand the social interactions and social dynamics taking place during the decision-making practice of funding committees are complex, and cannot be understood by examining peer reviewer or committee scores alone.^(11)^ Gallo *et al.* conducted a survey with a cohort of biomedical scientists to try and address the gap in the evidence by looking at the influence, quality, and effectiveness of their most recent panel meeting experience (e.g., being either teleconferencing or face-to-face panels). Although some panel members felt there was an unequal focus and limited engagement from unassigned panel members reviewing the research applications, which could lead to or limit the discussion on scoring, and possibly introduce bias, panel meeting discussions were viewed favourably (e.g., in terms of quality and effectiveness) and were perceived to facilitate the recommended funding decision.^(6, 11)^ However, a limitation to Gallo *et al.* study was that it only included a survey examining written/text responses. There were no observations of the panel meeting to confirm the individual responses from the survey.

To contextualise and understand the more subtle and implicit social interactions of funding committee practices, the exploration through surveys or interview methods alone may not be sufficient. Nonverbal cues provide additional meaning and observing the interactions (along with written notes) provides a more coherent and in-depth account of the social and interactional processes at work during online community settings such as funding committee meetings.

### Virtual funding committees and the role of netnography

There are a range of approaches used to conceptualise and understand the virtual social environment we now live in such as virtual ethnography, online ethnography, digital ethnography, and cyber ethnography.^(12–14)^ What distinguishes netnography from these forms of ethnography is how the research conducted follows a set of defined research tools, using a pragmatic approach, to study the cultural context and contents, including social dynamics, of online communicative acts in a virtual setting. ^(15)^ ^(13, 16)^

The National Institute for Health and Care Research (NIHR), the UK’s largest funders of health and social care research, uses funding committees to evaluate research applications and reach a consensus on the research to be recommended for funding. During the COVID-19 pandemic the NIHR ran virtual funding committees in place of face-to-face meetings for their research programmes. Following and using the principles of netnography, we explored, reflected, and investigated the new and changing landscape of NHR funding committee practice (virtual meetings).

The aims of the study were to explore virtual funding committee meetings in terms of the formal processes such as technology, resources and formality, and the informal processes such as the social interactions, social dynamics, perceptions, attitudes, and expectations. This paper describes a netnographic study on virtual funding committee practices to gain insight into using online forms of communications (e.g., cultural changes), the benefits, challenges, and barriers to using online platforms (e.g., future considerations) and understand the social interactions in virtual settings (e.g., members participation).

## METHODS

### Approach and study design

Due to the delicate nature of funding committee meetings and the confidentiality around the discussions and not attributing feedback to an individual committee member, netnography was particularly suited to answer the research questions. The methodological approach offered insights into the cultural processes in a virtual space that would not otherwise have been possible in a face-to-face setting. Netnography allows you to observe in an unobtrusive and non-invasive way (e.g., no observer presence is required), and although netnography shares similar foundations, perspectives and practices to ethnography, there are distinct differences in term of research focus, research methods, data collection and analysis.^(13, 15, 17)^ Exploring the nature and implications of the interrelationship between online social experience and how individuals alter in response to these new technologies is the foundation for netnography.

Netnography follows several fundamental stages like other qualitative methodological approaches, that are inclusive of

1. **research inquiry** (developing and initiating the research topic and approaches to formulate the research questions)
2. **collecting the data** (gathering the data through observations, surveys, interviews, online mechanisms, and through an immersive (self-reflective) journal)
3. **analysing and interpreting the data** (ongoing process of decoding, translating, and coding parts and segments of the data to seek narrative and thematic analysis)
4. **sharing the research** (contextualising and presenting findings in an appropriate form to disseminate the outcomes to the audience it was intended for).

Following interactionist principles, it was possible to explore the virtual conversations about how, where and when things were said, from the committee members through to the role of the chair in steering and managing the discussions. This was important for understanding how virtual social interaction and social encounters are different from physically embedded, face-to-face encounters.

### Research inquiry

To address the aims and objectives of virtual funding committees in terms of technology, resources, social interaction, social dynamics, attitudes, perceptions, and expectations we proposed to answer the following research questions:

1. How do virtual funding committee meetings provide an alternative approach for the recommended allocation of research funding?
2. Was there any impact of virtual funding committee meetings on the decision-making recommendations for research funding?
3. What were the key components and considerations of running and taking part in a virtual funding committee meeting and do they affect members’ experience?
4. How has the use of virtual online technology affected the social identity aspects of funding committee meetings?
5. Were there behavioural, attitude and relationship considerations (and constraints) when conducting virtual funding committee meetings?

#### Sources of information

To allow for divergent and in-depth interpretation of the online virtual funding committees observation, interviews and a survey were conducted following the guiding principles of netnography.^(13, 15)^ The interviews and survey were conducted after the funding committee meeting had taken place and the observational material had been obtained. The sources of information were complementary in nature and enabled cross validation of the observational data.

##### Observations

The virtual funding committees were recorded on the online platform used by NIHR staff to enable and assist in the minutes of the meeting outcomes which are made publicly available on the NIHR website. These recordings were used for observational purposes only and formed the basis of the netnographic study. We aimed to recruit between 8 to 12 funding committees, one committee meeting per NIHR research programme and funding committees were purposefully selected based on availability and programme engagement, commitment, and approval from the Programme Director.

##### Interviews

We aimed to conduct 25-30 interviews with funding committee members to understand their experience and expectations of a virtual funding committee meeting, and 10-15 interviews with NIHR staff to explore the practical challenges and potential benefits of virtual funding committee meetings. Several factors influenced the number of interviews needed to reach saturation, and methodologically, there is no definitive number to determine when ‘enough is enough’.^(18–20)^ Research by Guest *et al.* (2006) and Hagaman and Wutich (2016) suggested a range of 30 to 60 interviews for ethnographic studies.^(21, 22)^ We aimed to conduct a total of 35-45 interviews or until saturation was reached (e.g., reoccurring conversations with respondents did not emerge any new themes and sufficient data were retrieved to address the research questions).

A purposive sample was used to select funding committee members and NIHR staff based on the NIHR research programmes and on the role performed at the funding committee meeting (e.g., chair, clinician, methodologist, health economist and public representative) to ensure breadth of perspective.^(23)^ Invitations were sent to committee members and NIHR staff who attended the funding committee and all committee members were also invited to show their interest in being interviewed as part of the online survey. The interviews were recorded for audio and text data purposes only.

##### Survey

The survey was sent to all funding committee members included in the observational cohort to gain further insight and understanding of funding committee practice. A link to the survey (including online consent) to participate was sent to all committee members within four weeks of the virtual funding committee meeting taking place. Committee members were given three weeks to respond to the survey, with a two-week reminder, followed by a final reminder three days before the closure of the survey. We aimed to receive a range of between 160 to 240 responses based on the average size of 20 committee members per research programme. The survey was open for each research programme to participate during the period from October 2020 to January 2022 (based on when the committee meeting took place).

##### Documentary analysis

All materials provided to funding committee members were collected for analysis and provided a rich source of written text data to complement the material obtained from the online video footage, interviews, and survey. These documents included the agenda, chair’s brief, guidance for committee members including duties of members, funding committee roles and any After Action Reviews (AAR).

##### Immersive journal

The immersive journal was used to capture reflections, reactions, perceptions and meanings throughout data collection and during data analysis.^(15)^ This type of journal writing reflects on the process of doing the research, exploring new ideas, contextualising the data, capturing experiences, and providing extensive detail into the fragments of data. Immersive journals often contain the combination of what was seen but also what the individual experiences. Capturing these reflections allowed the research team to keep a record and provide any provisional thoughts for wider team discussion (see quality assurance section).

#### Identification and community sampling selection

All NIHR research programme funding committees conducted during October 2020 to December 2021 were eligible to participate in the study, including, Artificial Intelligence in Health and Care Award (AI award), Efficacy and Mechanism Evaluation (EME), Evidence Synthesis (ES), Global Health Research (GHR), Health and Social Care Delivery Research (HSDR) (formerly known as Health Service and Delivery Research (HS&DR)), Health Technology Assessment (HTA), Programme Grants for Applied Research (PGfAR), Public Health Research (PHR), and Research for Patient Benefit (RfPB). Each funding committee was classified as a single online community, based on its activity, interaction, size, and research focus.

#### Piloting

The netnographic study involved several methods and these were designed, developed, and piloted with a small sample to ensure their appropriateness. The sample consisted of NIHR staff from the application and funding teams, NIHR Patient and Public Involvement and Engagement (PPIE), programme chairs from the NIHR research programmes, and members of the research team. Particular attention was paid to the observation framework used to facilitate the pre-recorded online video footage from the funding committee meetings, questions in the online survey and the interview schedule. Modifications to the pilot were documented as part of the learning process (e.g., research focus and data collection).

### Data collection and ethical process

The study was approved by the University of Southampton, Faculty of Medicine Ethics Committee (ID 57541, November 2020).

#### Observations

All NIHR funding committees conducted during October 2020 to December 2021 were invited to take part. Only NIHR funding committees that had approval and agreement from Programme Directors and Chairs were included. Pre-emptive opt-out was used for the approval of using the video footage for research purposes only. This was explained in a covering letter accompanying the Participant Information Sheet (PIS) and was sent to the funding committee members and staff two weeks prior to the meeting, or at a convenient time agreed with NIHR staff. Attendees of the meeting had five working days to consider the option to opt-out of the observational study. There were no opt-outs (**see supporting materials S1** Appendix: Observation guide).

Observations of the funding committee meetings were first viewed for immersive purposes only to allow for personal reflections and initial impressions. This was followed up by a more semi-structured process, focusing on key elements noted in the observation schedule paying particular attention to the processes and practice of using virtual online technology as they emerged (and importantly related to the research questions). A passive-observer position was taken (the research team was not present) as this was the most unobtrusive research approach. The online funding committee meeting recordings were deleted once they were analysed.

#### Interviews

Two interview guides were used for the two groups of participants: funding committee members and NIHR staff (**see supporting materials S2 Appendix: Interview guide for committee members; S3 Appendix: Interview guide for NIHR staff**). Where possible, interviews with NIHR staff took take place within a week of the committee meeting, and interviews with the funding committee members were conducted in parallel with the online survey. The participants were purposively selected and invited from pre-defined lists (using Microsoft Excel random number generator), sorted by the relevant NIHR programme. Identified committee members and NIHR staff were sent an invitation letter along with the PIS. They were given two weeks to respond, and a reminder email was sent out after one week. If they expressed an interest in participating, they were contacted to discuss the study requirements and a date was arranged to conduct the interview. Where there was a non-response from the invitation, another set of participants were randomly chosen until we had a sufficient number of committee members to interview.

Any committee member who completed the online survey and expressed an interest to be interviewed, was contacted, and included in the study. This enabled greater flexibility and inclusivity for those who may have had additional experiences to share with the research team. The interviews took between 20-60 minutes, with an opportunity for the participant to follow up on any additional points not covered in the survey or interview schedule. Semi-structured, open-ended questions with prompts were used to inform the interviews. NIHR staff and funding committee member interviews followed the same structure although the focus and topics of interest varied.

The interviews were conducted using Microsoft Teams, or if this was not feasible due to international location or internet connection, Google Hangout and WhatsApp platforms were used. Research data was recorded in the form of audio and visual files where applicable. Verbal consent was gained from all participants prior to conducting the online interviews. None of the interviews were transcribed and the interview recordings were deleted once they were listened to, and notes were taken (and as part of the immersive journal).

#### Survey

The survey for funding committee members was sent within four weeks of the virtual funding committee meeting and included closed and open-ended questions and Likert scale responses (a total of 16 questions, with 5 follow up questions) (**see supporting materials S4 Appendix: Survey questions**). The participants were given three weeks to respond to the survey with two follow up reminders (a two-week reminder, followed by a further reminder three days before the closure of the survey). We anticipated the online survey would take approximately 15-20 minutes. Online consent was required from all who completed the survey. The online survey was hosted on a University of Southampton server and participants could access the survey from anywhere that had an internet connection.

### Data analysis and interpretation

As the study included several methods and approaches, these were drawn on to analyse and interpret the concepts and constructs of virtual funding committees. Both qualitative and quantitative data arising from the study were complementary in nature (rather than competitively) and integrated analytical and interpretative data operations simultaneously.^(13, 15, 24)^

All qualitative materials (including text data from the virtual observations) were analysed using an inductive approach, allowing the data to drive the thematic coding. Microsoft Excel and Nvivo software were used to analyse the data, where appropriate. Online survey results were downloaded and initially analysed using a Microsoft Excel spreadsheet, which were later imported into Nvivo to enable cross validation with the observational and interview data.

The authors independently analysed the data collected from the observations, interviews, and open-ended questions from the survey, which informed the development of the initial themes. The themes were categorised, analysed, and compared across the three data collection approaches. The initial coding and categorisation of the data provided key headlines to help establish and develop the main themes. Within each of the main themes, sub-themes were used to represent the range of topics that were extracted across the three main data sources (observations, interviews, and online survey). The themes and categorisations were independently extracted for each of the data collection methods, and then reviewed by the research team to determine where there were commonalities between what was observed, what was spoken through interviews and what was reported in the online survey. The teams’ immersive journaling also formed part of the verification steps and consensus on emerging themes. Translating the data and seeking consensus and agreement from the research team took place simultaneously during data collection and amendments to categorisations or themes were recorded in Nvivo for transparency purposes.

#### Quality assurance

In all qualitative research there is a question around the potential for researcher bias. Due to the research team’s experience and background in the allocation of research funding from the NIHR, there was the potential for preconceptions and biases to occur. To minimise researcher bias, there was more than one researcher on each type of data collection (e.g., interviews, online survey, and observations) to either double code or to review and discuss the preliminary analysis. The research team was also encouraged to keep an immersive journal noting down any reactions, feelings, and readings from the observations, which were used to discuss different perspectives and understand any potential unconscious biases. The research team was therefore confident that this helped to minimise individual and group bias. This was led by the lead researcher to ensure continuity across the study.

For data processing, several approaches were used to not only process the data but also to maintain quality assurance measures of the collected data in the study. The large volume of data consisted of audio, videos, transcripts, text data, immersive journal notes and survey data, which were imported and held in Nvivo software and Microsoft Excel. Combining the data allowed for more divergent thinking and allowed for meaningful interpretation from different sources of data collection, including the verification and validation of the research claims from the observational data. Collecting data using different approaches also allowed for greater interpretation that would not otherwise be possible from the observational data alone.

## RESULTS

A total of six of ten NIHR funding programmes agreed to take part in the study and nine funding committee meetings during the period of October 2020 to December 2021 were included in the study. Having two NIHR programmes include more than one meeting provided the opportunity to explore variations over time through the observations but also through the follow up interviews. The online survey was active from October 2020 and closed four weeks after the final committee meeting (January 2022). This allowed all committee members from all committees to participate in the survey during the four-week timeframe after the committee meeting took place. Fifty responses were received from a total of 222 invited respondents (response rate of 22.5%). An invitation to participate in an interview was sent to 60 committee members. Eighteen interviews were conducted with funding committee members across the nine funding committees and included a range of committee roles such as public contributors, statisticians, health economists, clinicians, funding committee Chairs and NIHR programme directors (response rate of 26.6% (16/60) from the invitation to participate, and two from self-selection from the online survey). Twelve interviews were conducted with NIHR staff who participated in one of the nine funding committees and included senior research managers, research managers and assistant research managers (response rate of 44.4%, 27 invitations were sent to NIHR staff).

The demographic characteristics of the participants involved in the study and the online survey were not collected due to confidentiality. There was wide coverage across all groups of committee members ranging from patient and public representatives to health economists. To further prevent the possibility of individual exposure a high cloaking level was taken across all forms of data analysis and verbatim quotes were amended to remove any associations with funding committee members. All quotes (written and verbal), used from the three sources of data collection, were therefore labelled as P1 (survey), P2 (interview) and P3 (observations).

### Emergent themes

Three main themes, each with two or three subthemes, were extracted across the three main data sources (observations, interviews, and online survey). Data also revealed a range of experiences between the NIHR funding programmes, but this was not explored further due to potential exposure of participating committee members. **Fig 1** illustrates the three themes followed by the sub-themes. It is important to note that it was common for participants to make comparisons with face-to-face funding committee meetings during the interviews and survey, and whilst these comparisons are reported they were not observed directly.

**Fig 1:**
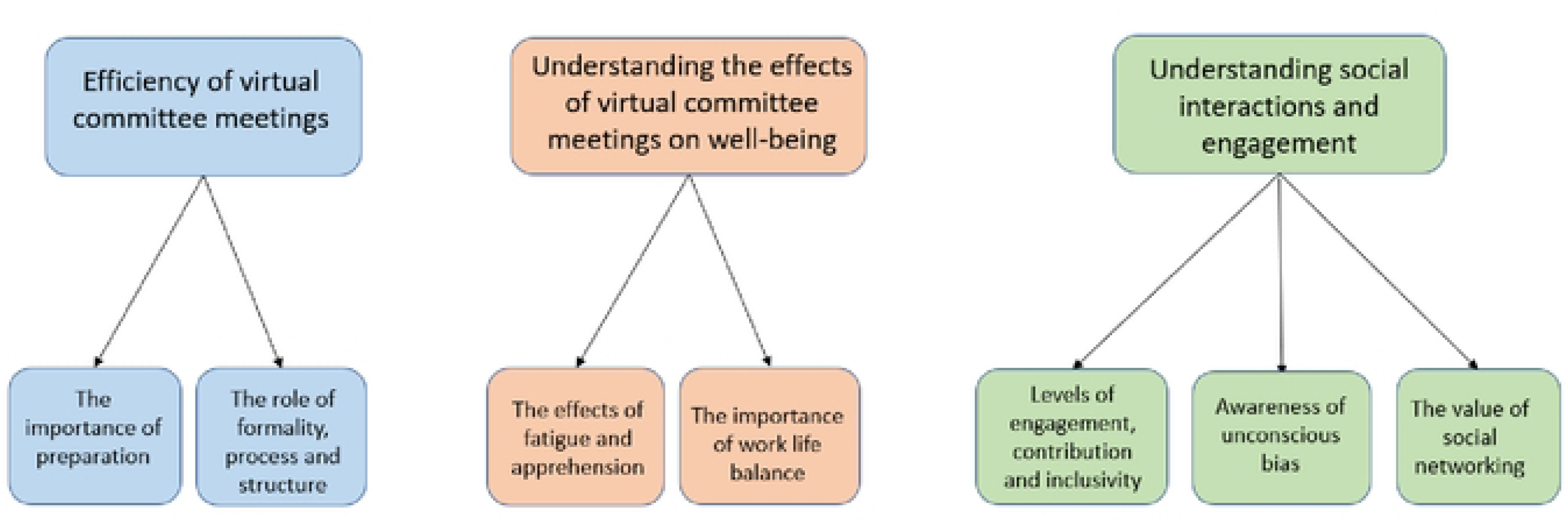
Emergent themes and sub-themes

### ADD Fig 1. HERE

#### 1. Efficiency of virtual committee meeting

The function and structure of virtual funding committee meetings were a key consideration for all respondents, particularly around the duration of the meeting and the effort required to prepare and run these meetings virtually (particularly for NIHR staff and the Chair). A majority of respondents (37/50, 74%) felt that you could achieve the same outcome through online virtual funding committees compared to face-to-face meetings, and 94% (47/50) of respondents felt that virtual committee meetings have a role to play for the future allocation of research funding (**Table 1**).

> *“Yes, most of the process is the same. You can still read the projects beforehand and score them as it was done before. The discussion is the same and the decision can still be made in the same way using technology (e.g., to vote).” (P1)*

**Table 1.** Responses to whether respondents saw any potential opportunities, benefits, barriers, or challenges to using virtual online platforms to allocate research funding.

| Survey responses | Yes | No | Did not respond | Total |
| --- | --- | --- | --- | --- |
| Survey questions |  |  |  |  |
| Do you see any potential opportunities and / or benefits to using virtual online platforms to allocate research funding? | 43 (86%) | 4 (8%) | 3 (6%) | 50 (100%) |
| Do you see any potential barriers and/or challenges to using virtual online platforms to allocate research funding? | 36 (72%) | 12 (24%) | 2 (4%) | 50 (100%) |
| Do you feel you can achieve the same outcome through online virtual funding committees compared to face-to-face meetings? | 37 (74%) | 13 (26%) | 0 | 50 (100%) |
| Do you feel that conducting funding committees virtually | 47 (94%) | 3 (6%) | 0 | 50 (100%) |
| have a role to play for the future allocation of research funding? |  |  |  |  |

> *“I’ve now attended two virtual meetings and they went much better than I expected. I had worried my participation would be negatively affected from being on a screen all day but that doesn’t appear to have happened. I feel that I’ve been able to contribute fully as if it were an in-person meeting.” (P1)*

In addition, although 86% (43/50) of respondents felt that there were potential opportunities and/or benefits to using virtual online platforms to allocate research funding, 72% (36/50) also felt that there were potential barriers and/or challenges (**see Table 1**). Thus, funding organisations need to carefully consider the potential trade-offs of conducting funding committee meetings virtually.

Included in the benefits of virtual meetings, multiple respondents highlighted the cost and environmental benefits to running these committees virtually and this was a strong motivator across respondents for using virtual platforms for committee meetings in the future. For example, *“efficient use of time, travel, environmentally more friendly – better for inclusivity and diversity.” (P2*) By contrast, fatigue and social disconnectedness was frequently mentioned by both committee members and NIHR staff as the main challenges with running these committee meetings virtually. For example, *“Of all of those fears - gave it a thorough test out, it worked really well. There was no residual doubts. It does work, it’s a different challenge…some of the anxieties are unfounded. It’s physically and mentally exhausting…mentally more demanding.” (P2)*

##### The importance of preparation for virtual committee meetings

The level of appreciation for the preparatory work in coordinating virtual committee meetings was noticeable by all committee members and NIHR staff. Having reservations about conducting committee meetings virtually were a concern for committee members (including the chairs), which resulted in several preparatory steps by NIHR staff to support attendees and alleviate any known anxieties.

> *“You’ve got to really really sing the praises of the secretariat here I mean, they gave it a huge amount of thought beforehand and they tested it out and they made sure me and XX and other senior members were prepped […] it was a real testament to them, the secretariat, that they anticipated what the problems were and simplified things to a point that busy people could interact and they didn’t stick to what was convenient to them. They took on board the kind of feedback and simplified it.” (P2)*

As noted by the NIHR staff there was a tremendous amount of work undertaken prior to the meeting, such as testing, piloting, carrying out pre-runs of the committee meetings, working out how to use the virtual platforms (for conflict of interest on applications) and voting systems. The amount and level of prior preparation for virtual committee meetings was frequently mentioned by NIHR staff and all eventualities were considered to try and prevent any delays during the actual committee meeting.

> *“We do one re-run to make sure everything works together. Use Teams, but the presentation is on a different package, voting is on another. So we need to make sure everything works together to run but also be in sync with one another. Lots of preparation required especially when there are updates to the platforms prior to meetings, so this is intense for the secretariat staff.” (P2)*

For some NIHR programmes, there was an added level of complexity, as some committees had new members (or were a new committee group) and some committees included international members which meant time zone differences needed to be considered during the preparation. To accommodate new membership, the NIHR staff prepared and run induction and introductory sessions prior to the committee meeting, so that members were familiar with each other and how the committee meetings are run. The chairs were also involved in these sessions, as there were also new chairs and deputy chairs running the committee meetings as well as conducting them virtually. These challenges can have an additional impact on time keeping and trying to maintain the order of the agenda.

Although, the set up and preparation involved more staff and more time, there was general consensus that it worked well and met the needs and purpose of running committee meetings: to make recommendations to fund research.

> *“The most important thing is quality and decisions, we have decades of doing it face-to-face, we understand that approach, we have a relatively limited, forced into it limited exposure to virtual. So far so good” (P2)*

> *“We are now seriously talking about now having more meetings as actually through choice making most of those meetings online so I think clearly if it hadn’t worked, we wouldn’t be having those conversations*. *So, I think that is a testament to how well it’s worked.” (P2)*

It was clear that virtual committee meetings are conducted differently to face-to-face meetings for several reasons, which need to be considered if virtual platforms are considered in the future. For example, one interviewee commented that virtual meetings were “*less free flowing and more structured*.” However, it is important to note that NIHR staff followed the same structural format in terms of paperwork, reviewing, preparation and decision-making. The process to decision-making didn’t change, only that it was conducted online rather than in a room face-to-face.

##### The role of formality, process, and structure of virtual committee meetings

Providing clear structure at the beginning of the committee meeting helped to prepare committee members on what was required of them, the order of the discussions and to ensure all attendees contributed to the discussion. The intensity and increased number of staff required to run these meetings virtually was recognised by both committee members and NIHR staff. For all committee meetings, more NIHR staff were required to ensure every aspect of the meetings ran smoothly, and in some instances NIHR staff also had to manage separate online chats to coordinate and manage the running of the committee meeting on Microsoft Teams or Zoom. However, similar to preparation for meetings, the challenges of virtual meetings became less as more experience was gained from conducting these committee meetings virtually. Several respondents highlighted how the process and structure became easier second time round, as they knew what they were doing.

> *“Now we are in the flow of the meetings, it’s the new normal and that’s the way it is and we kinda know what we are doing and we have had to adapt to it. But we are used to it and it works well, I think a lot of colleagues prefer it this way. There are no other superficial problems to deal with such as the venue with food or a room or the air con. It simplifies the meeting in a lot of ways.” (P2)*

It was noticeable in all committee meetings that there were clear boundaries, guidance and expectations set out at the beginning of the day, to ensure that all members understood how the day was going to be run. Setting out how the meeting was going to be handled was inclusive, with the committees spending time introducing themselves and encouraging members to turn their cameras on when discussing an application or wishing to participate in the discussion. Although this took time out from the committee discussions, it was appreciated by committee members, especially when there were new members or a whole new committee. However, it was also notable that committees who were more established and known to each other, the flow and structure appeared less formal. This did not deter away from the purpose of the committee meetings, rather it provided a more relaxed atmosphere at a very intense and demanding time. There were also notable differences to the formality of the discussions based on the size of the committee, which did affect the running of the meeting virtually in terms of technology and ability to have cameras on for a committee size of seven compared to 29.

> *“I did wonder about new committee members joining […] so at the moment we got a committee that knew how the face to face ran and know each other, and how that might play into things moving forward making sure that people are supported coming into this format if it continues.” (P2)*

Almost all committees used the Vevox system to produce the final scores on each application (identical system used in face-to-face committee meetings). The use of the software was extensively tested prior to the committee meeting, and all committees tested the software at the beginning of the meeting (e.g., all members could access and cast their vote, and the results could be presented on screen for committee members). Although there were some glitches across all committee meetings, with varying ways to resolve these issues, it did not appear to impact the overall decision-making for funding recommendations. All chairs provided clear guidance around the threshold scoring, which was universal across all committee meetings.

For most committee members, the voting system was familiar to members, so despite some technical issues, respondents were encouraging about its use. For those that did not use Vevox for scoring, other methods included using an excel spreadsheet and individual committee members scores being vocally given at the end of the application. The size of the committee and how well established they are was factored in during the preparatory work for the committee meeting and to determine the appropriate approach for scoring. All the scores, for all programmes were collated by an NIHR staff member and all applications when ranked were discussed.

Conflict of interests were given adequate time and attention during the preparation of the meeting, particularly for the order of applications. This becomes even more important when running committee meetings virtually but even more so when committees included members from different geographical locations (e.g., international locations). Unlike face-to-face meetings, where committee members leave the room, this is more complex to manage virtually. A similar process to deal with conflict of interests was used by all committees, although there were variations between different virtual platforms due to the functionality options available at the time. Over time, this also meant virtual platform updates enabled more options for NIHR staff and the committee to consider as part of the planning prior to the committee meeting.

> *“So, whatever I am saying now is based on having run it twice, whereas things may change, the software may change we have already found between the first and second one that some of the functionalities are improving the more we do these remote…we are just running them remotely as general working.” (P2)*

For some committees it was challenging to manage the conflict of interests, which meant that the NIHR staff needed to be fully engaged to ensure all those with conflicts did not return to the meeting until the scoring outcome was removed. Conflict of interests were a particular concern if comments were written in the chat function as everybody, including those who had left the meeting, can see the comments made. Decisions were made early on by the NIHR staff, that the chat functionality was not to be used for comments, only to raise a hand, inform the committee they were stepping away from the meeting, or would like to contribute to the discussion. Over time, there were adaptations to the process and some committees reviewed the process and made relevant changes to accommodate members and make it more streamlined.

> *“It is trying to make the whole experience easier and better for them in every round that we do Some committees are more adept to changes than others, it’s not that they are resistant to it, it’s just the nature of the committee and they are not able to move as quickly as some of the other committees. We have to be mindful of that as well.” (P2)*

As mentioned by respondents, the chairs’ role is challenging and requires different skills in the virtual environment. Different chairs had different styles of chairing and contributed to different levels in the discussion. However, all chairs frequently reminded committee members about the review and decision-making process of the committee, enabling constructive feedback around what was typically referred to as ‘fixable flaws’, ‘fixable faults’ or ‘fundamental flaws’, “*could you go through any fundamental issues and anything that could be fixable please?” (P3).* Whilst sometimes the transition between chairs wasn’t always smooth, each chair encouraged all members to contribute to discussion. It was suggested and observed that the quality of the chairing made the discussions what they were.

> *“Chairing these things effectively - you don’t know how difficult it is…a good chair makes it look easy…chairing is hard work.” (P2)*

> *“Good chairing helps to ensure that people are able to ask questions and make comments, although discussion dynamics are much easier in person, when you can see everyone in the room.” (P1)*

#### 2. Understanding the effects of virtual committee meetings on well-being

As raised in theme one, adopting and changing a critical part of the decision-making process for the allocation of funding due to the COVID-19 pandemic has allowed for alternative approaches to be used. Most respondents have adopted and embraced the changes however, it is important to understand that not everybody is comfortable with virtual environments, and the effects of virtual meetings on well-being raises important considerations around the future recruitment and retainment of committee members. Committees are made up of a diverse group of individuals, and what may work for some may not work for others, and this can have important repercussions for committee members not feeling fully inclusive or equal to other members of the committee.

> *“Given that it worked, and I suspect that it will continue to work, I think it’s really a different style of solicitation of views, quite a few people could find it easier, Designated Committee Members find it easier talking into a machine. Could be easier for some but harder for others.” (P2)*

##### The effects of fatigue and apprehension

In earlier committee meetings, there was apprehension about how it was going to work, and it required extensive preparation and resourcing to try and have a plan for every eventuality. As a result, the planning and preparation, alongside attempts to resolve any uneasiness from committee members, resulted in fatigue for NIHR staff.

> *“We had worked out a way for delivering them face-to-face and therefore there was some anxiety when we were forced to do them online, it was either online or not do them and not doing them was just incomprehensible, you just couldn’t work out what would happen if we didn’t do them.” (P2)*

This was also coupled with the fact that the meetings were conducted over two or three long days, and often over ran which meant reducing the scheduled break time or continuing until after the proposed finish time. All participants (NIHR staff and committee members) acknowledged that these meetings were always intense and required significant work, however, it was felt that participation and contribution to virtual committee meetings presented different challenges and issues than those experienced at face-to-face meetings.

> *“It is tiring working online for two consecutive days and one is less able to concentrate to the same degree and over the same period of time as in the in-person meetings. It is easier to disengage.” (P2)*

> *“As chair I found it physically and mentally exhausting - more than face-to-face – it’s more demanding online - as you are not getting all of the visual cues. It’s not the same thing, some are out of focus, poor signal.” (P2)*

For example, respondents highlighted several key concerns such as eye strain from staring at a screen all day, back pain from sitting looking at a screen in one position for longer, and mental fatigue from additional demands in keeping up with conversations online. These issues often made it challenging to be attentive for the whole duration of the meeting.

> *“The agenda needed to be more realistic, with more time for breaks. I had serious screen fatigue!” (P1)*

Due to this, some participants reported that virtual meetings had more challenges and longer-term implications for their wellbeing. For others, virtual meetings were helpful, and for one participant who was hard of hearing commented:

> *One issue that is surprisingly better for me - against expectations – […] If you can’t hear someone on a virtual meeting you just say speak up and people do, automatically! (P1)*

Another compounding factor on tiredness and fatigue was the duration and frequency of breaks. Although NIHR staff built in breaks and tried to allow for more breaks, compared to face-to-face meetings, they did not always happen as often the virtual meetings were over-running and so breaks were sometimes delayed or cut short. In addition, participants commented that breaks in virtual meetings are different from those experienced at face-to-face meetings. For example, at face-to-face meetings the drink and food is prepared for you, whereas in virtual meetings participants had to factor making food or drinks into the break time. It was felt that this resulted in less free time or break from their screens.

> *“…it’s a long time to concentrate. You have to prepare your own food, so break time is down time to quickly prepare food and run back to your desk. It’s harder in that respect, as longer breaks would be better.” (P2)*

Some committee members also felt that they got more breaks in face-to-face meetings as often they would have to step out of the meeting because of conflicts of interest. In the virtual environment, participants reported that they could never step too far away from the computer as they were not sure when they might be called back in again and so were always on alert.

##### The importance of work life balance

All respondents appreciated there were benefits and challenges associated with virtual committee meetings. For most it was welcomed due to not having to travel and stay overnight, no early mornings or late evenings, taking additional time away from other work or family commitments, ease of participation and more efficient use of time without jeopardising the process.

> *“…you can sleep in your own bed, you don’t have a thousand-mile round trip, because that is tiring in its own right.” (P2)*

> *“Not having to get up at 4am to make it to London for a meeting and being tired all day. Not having to be away from family and my usual routines. Being able to exercise and eat properly at home and follow my usual routines. Being more relaxed when it wasn’t “my turn”, but still being able to contribute well, listen effectively, and vote.” (P1)*

Thus, for some, virtual meetings offered greater flexibility to manage a work-life balance, and many respondents indicated that this better accessibility might encourage and/or facilitate more people to become committee members and allow for more diverse membership. However, given the need to move to virtual meetings because of the pandemic, challenges due to all schools and colleges being closed and other restrictions on activities, resulted in additional complexities and requirements during home working. From observing the committees, all members and NIHR staff were sympathetic to the demands placed on individuals and there was regular commentary from the chairs.

> *“…what a challenging time we are living in…well done for balancing childcare”* Or *“…let the committee know if you have to leave by using the chat as we, the secretariat appreciate that working from home means that some of us will have personal commitments to deal with.” (P3)*

There were also comments about virtual platforms allowing a glimpse into colleagues lives that might not otherwise be shared. Although these comments were said in a positive way and participants enjoyed seeing personal backgrounds and pets, for some it added to the blurring of home and work life which they preferred to keep more distinct.

> *“…one of the nice things about virtual are seeing other people’s backgrounds - is fascinating; and produced a new angle on getting to know people.” (P2)*

#### 3. Understanding social interactions and engagement

##### Level of engagement, contribution, and inclusivity

Throughout the observations and responses from the survey and follow up interviews with respondents, it was evident that engagement, inclusivity of members, and contribution to the discussions were prominent areas of consideration for virtual committee meetings.

> *“Structure and pace of the meeting was excellent, and it was very well chaired. Keeping to time and encouraging contributions from all members of the committee.” (P1)*

Although for many virtual meetings were not a new concept, for some there were practical challenges associated to having the committee meetings virtually. This was particularly relevant for those that did not have the space or sufficient computer equipment such as having two screen monitors or a good internet connection. These issues were seen as being disruptive and problematic, and meant engagement and conversation was at times challenging.

> *“Very much dependent on internet - so when there is a delay it can feel awkward, especially for voting.Some people do not like the long silences and find it hard to cope with.” (P2)*

The committee meetings observed showed that there was good quality discussion with input from a variety of members (and equal opportunities provided to contribute to the discussion), providing diverse discussion and indicating member engagement. However, judging the engagement levels in virtual meetings is difficult, especially if someone has their camera off. Indeed, there were challenges around having cameras on for some committees and although NIHR staff and the chair tried to encourage members to have their cameras on whilst presenting or joining the conservation, there were technological issues associated to this.

Those that were on camera were not always looking at the screen. Although some committee members indicated that this was to reduce eye strain or because they were looking at another screen or writing notes, this sometimes gave the impression that they were not engaged with the discussion. Interviews and survey responses also indicated that committee members felt it was harder to concentrate for long periods of time during virtual meetings and they had more distractions to contend with in the home setting.

> “…*it’s mentally more demanding…it’s more difficult to keep the conservation going…in a room you can pick up on the visual cues, you can’t do that online.” (P2)*

> *“If in face to face you know that you are committing a whole entire day to be there so out of office is on, don’t check emails etc BUT in virtual you can get distracted by emails more easily; virtual gives you flexibility to switch on and off and so may lose concentration.” (P2)*

Furthermore, because of the virtual environment, some members were at work or had just arrived home from a day’s work (this was particularly true for non-European countries and the time zone variations). Although this demonstrated inclusivity and allowed diversity of membership, this added an extra layer of distraction and fatigue to these members which may have had an impact on their level of engagement during the committee meeting. For example, attendance at meetings with international members was often found to drop off as the day went on due to different time-zones.

Having cameras off was found to make it difficult to read social cues and body language. The value and importance of ‘reading the room’ for some was just as important as the conversation itself, and some felt that this social connectedness was totally missing through virtual platforms.

> *“You do have some visual cues online but it’s just not the same as being in a room, the visual cues are just different. The quality of the cameras vary, the connections vary.” (P2)*

> *“When you’re in a room there is a way to negotiate time, in a sort of untold way by using visual cues and looks and all kinds of things you can do to create your space. This is missing completely*.” *(P2)*

This was found to be particularly challenging for chairs because it was harder to know if further discussion was needed or if the committee were generally happy with the decision. Chairs encouraged inclusivity but found it more difficult to ensure they were providing sufficient opportunity for all committee members to contribute to the discussion in a virtual setting due to the lack of social cues. This was particularly noted as the meeting progressed throughout the day. However, all chairs continuously engaged with the committee, openly providing members the opportunity to contribute, and therefore maintaining a quorum.

> *“Earlier proposals in the day received wider participation but by the end of the meeting, few people could contribute, understandable as it was a long day.” (P2)*

> *“The breaks were brief, and people were tiring and as the meeting went on more cameras went off and people were becoming less engaging.” (P2)*

> *“I am not sure how to improve the inertia towards the end of the meeting - understandably the members were tired, and the chair and secretariat did an amazing job of ensuring that the last proposals were also discussed in depth.”* (P2)

Chairs found it difficult to always spot when someone had something to say and how to bring different members into the discussion, noting that ‘*it is a lot harder than it looks’*. Different techniques were used between programmes and the level of support from NIHR staff varied by committee meeting. In line with this, some members commented that they felt it was more difficult to follow the conversation, to interrupt discussion and to add their contributions during the committee meeting.

*“Etiquette is that you keep camera off unless speaking so most chairing is done in the dark” (P2)*

> *“The introverts who are waiting to contribute and Teams does not do them justice. I am see that in a room but not online - it’s really hard…it’s a big committee and the noisy voices are heard more. So, I have to invite members to talk.” (P2)*

There were differences across the programmes on how long the Designated Committee Members (DCMs) spoke for about an application and what was included in their summaries. However, the purpose of stage 1 applications is different to stage 2, in that stage 1 applications are shorter, and the primary purpose of the committee is to assess the quality and value of the research question. For stage 2 applications, the role of the committee is to make a funding decision based on the full application. From the observations, at stage 1, decision making often relied more on committee member comments and there was less discussion. At stage 2, discussions were fairly varied as members with different roles were assigned as DCMs for each application – e.g., clinician, methodologist and patient and public contributor, so each reviewed the proposal with a different perspective. This resulted in some applications having less or more time allocated, as the chair typically but not exclusively would do a final summary of each application prior to the scoring of the application.

> *“Pace sometimes quite fast but generally sufficient discussion.” (P1)*
>
> *“…it is faster paced online, and you need to be quick to jump in.” (P2)*

Nevertheless, all data sources indicated that moving to virtual does not seem to have had an impact on the overall decision making. It was felt that the virtual setting made the discussion more focused and there was less deliberating over every point. It was also felt that contributions were made by different members, which indicated that committee members were engaged with the process.

> *“…think that it is highly unlikely that virtual impacts on the quality of the decision, as most of the preparation happens beforehand.” (P2)*

> *“From the quality of the contributions going remotely hasn’t had any impact in terms of engagement. Where I think it’s impossible to make a call is when the most interesting conversations happen when you push it out to the floor … it is infinitely more difficult to do that remotely.” (P2)*

##### Awareness of unconscious bias

Unconscious bias is a function of being human and so it is unlikely that any decision-making by a funding committee will be completely free from bias in whatever format committee meetings are conducted (virtual or face to face) but recognising such biases may be present and acting upon them is important.^(25,26)^

Unsurprisingly committee meetings with many members, some behaviours were observed in this case that could potentially reflect unconscious bias. All respondents were clear that it was essential that all applications were given a fair hearing, and overall, this is what was observed in the virtual context. However, as with face-to-face committees, when time was short, or the committees were over running, there were instances when the committee might not spend as long discussing applications with a clear trajectory of a very low or high score.

> *“Very positive comments indeed. External peer review comments are also very positive so don’t want to spend too long on this.” (P3)*

It was observed that a lot of emphasis was placed on external reviewers and DCMs (three or four committee members who were assigned to review a particular application) scores in the assessment process. This is usual practice, and there was no variation between the virtual setting to face-to-face. All committee members had the opportunity to contribute to discussions on an application. However, there were examples of some mismatch in the scores and written comments given (from the external reviewers and DCMs) and some clustering of scores. For example, written comments might highlight multiple and significant flaws in a study and a score of four is given (which is fundable) or lots of strengths of the application are given and then it wasn’t scored highly. The interviews highlighted that NIHR staff, and some committee members were aware of this issue, in that the committee scores were the only ones considered in the overall ranking of the applications. Pre committee scores can, however, change because of committee meeting discussions as other members raise issues on an application. Chairs would encourage committee members to use the range of scores available (1-6) and described what the numerical scores meant at the start of the committee meeting. This was repeated throughout the committee meetings to ensure balance across all applications.

It was also observed that how things were said and by whom can also contribute to potential unconscious bias by framing an application in a positive or negative light. For example, “*beautifully written*” or “*This is the first time I’ve given a 6; it’s very impressive, one of my favourites*” presented applications in a positive light, whereas “*eye wateringly expensive*”, or “*this one is a bit of a marmite application*” suggested room for improvement *(P3).* Some respondents expressed that they felt this happened more with experienced or senior members and in committees that had more established members rather than several new members of the committee.

> *“When certain people have opinions and say something in an authoritative tone it can become the mood of the room” (P2)*

> *“Perhaps one thing is the virtual space allows for the dominant characters to dominate…There are people who dominate more and it’s transferred to the virtual space. So, it’s finding ways to deal with this - mechanisms to ensure everyone has a voice using the virtual space as the platform for committee meetings.” (P2)*

There were some instances when committees paid particular attention to an applicant’s gender or career stage. For example, in a summary of an application one committee member noted, “*led by two female PIs.”* It was felt that this comment was meant as a positive one, by which committee members were paying particular attention to improve research equality, diversity, and inclusion. Another example was when a dedicated committee member stated when introducing an application “*this is one where there is a junior PI and I think they provide quite good justification for that.” (P3)*

Across all programmes, the observations found that some members of the committee were referred to by their role in the discussions. Although, often this was done in a factual way or because they needed certain expert advice (e.g., health economics), this was most notable for patient and public contributors. For some respondents, it was also felt that the larger the committee the more opportunities for bias could be introduced.

From the observations, it was noted that one NIHR committee was actively engaged in increasing awareness of unconscious bias through the use of a video training. In the second observation of this committee, members were observed to recognise unconscious bias on occasion indicating an increased awareness in this topic. There was also the discouragement of using the chat function as it could have been seen to introduce bias.

> *‘…this is probably my unconscious bias but…’ (P3)*

##### The value of social networking

One of the biggest challenges with conducting committee meetings virtually was the lack of social interaction and networking opportunities. The importance of social networking was frequency mentioned by members of the committee and NIHR staff. The survey data found that 54% (27/50) of respondents considered face to face networking at funding committees as very important to them and 34% (17/50) reporting it was somewhat important (**see Fig 2**).

**Fig 2.**
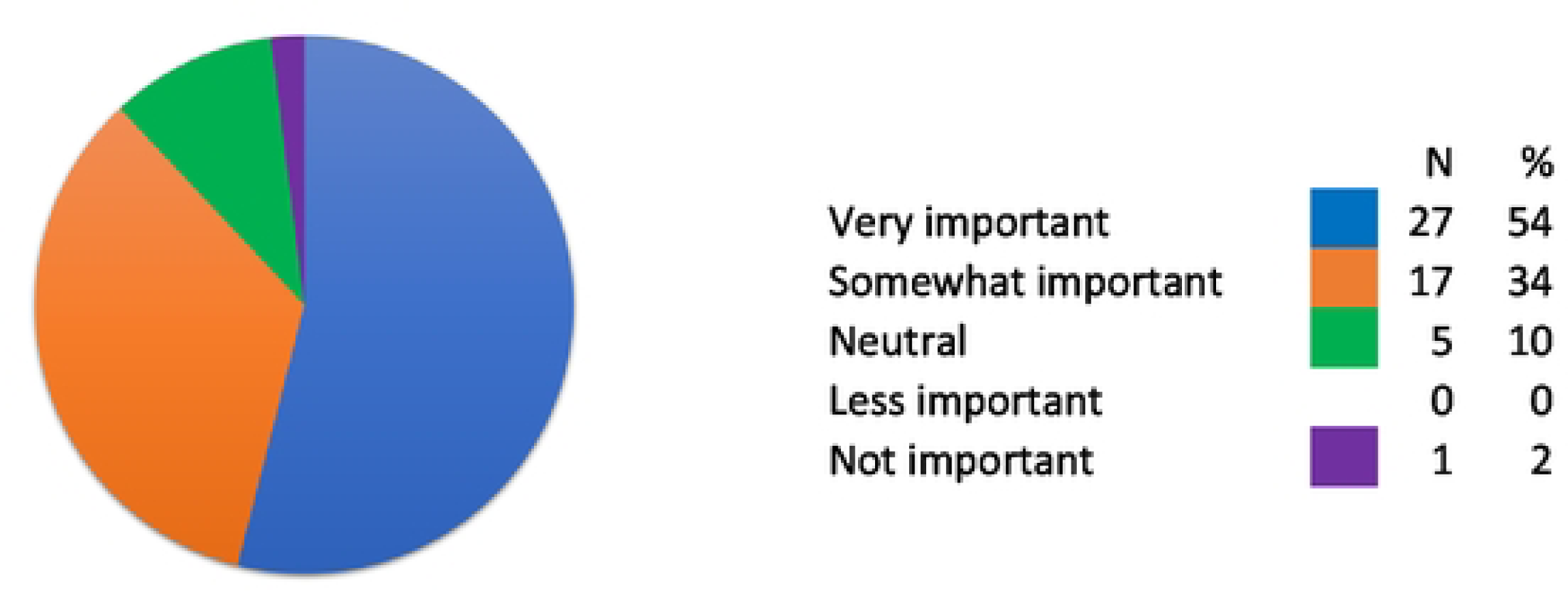
Responses to the importance of face-to-face networking at funding committees (N = 50)

### ADD Fig 2. HERE

A range of social aspects were described by respondents as lacking through having to conduct the committee meetings virtually, which were not entirely related to just the meeting itself. The survey reported that 66% (33/50) of respondents did not consider the additional features of using virtual platforms to conduct funding committees (e.g., chat function, raising your hand, and voting methods) provided more opportunities for members to engage in the committee discussions. Several committee members and NIHR staff also reported missing the chance to socialise during break time and the social gathering and networking after the committee meeting during virtual meetings.

> *“…we did set out what we wanted to achieve, it was three incredibly long days without the nice sort of nice relaxation at the end…you all go out for a nice meal and relax and talk to the committee members in a more relaxed and social environment. That social aspect was missed and although that social aspect is not vital it’s not what we are there for it oils the wheels.” (P2)*

Getting to know other committee members was suggested to help with understanding context for comments from certain members and it was evident that more established committees had more conversational dialogues and light-hearted comments.

> *“Miss out on the conversations at coffee time - discussions more around general things and helps us to understand where each other comes from. But you still learn from this. You’re able to ask questions for example, about a particular point or method. We can go and talk about it, about what do you think about this. So, it’s the whole, not just the yes no we approve an application, there is more to it than that and we miss out on that. It becomes very process driven, a tick box process in some respect.” (P2)*

Social networking was also suggested to be particularly important for new members to ‘get to know’ the other committee members. For some respondents, it is during these social interactions that collaborations and networks are formed, and this was seen as an important part of becoming a member of a committee. It was also seen as a positive benefit from spending what would ordinarily be non-work time with existing committee members, given the level of commitment required during funding committee meetings. Participants reported that this lack of social and networking engagement for future committee members could impact whether individuals choose to join a committee in the future. These opportunities provide new members with a sense of feeling included and key to being integrated as part of that committee and developing oneself at a personal level. Without these social interactions, for some respondents, there was a loss of a sense of belonging as joining a committee the first time was described as ‘daunting’.

> *“It is daunting when you’re a new member and social engagement and interaction is key to feeling integrated and part of the committee.” (P2)*

Several committees encouraged social networking as part of the preparation of the meeting, especially for those committees that were newly established, had several new members or involved several international members. This was received well by new members, although this added additional pressure to the preparation time for NIHR staff.

> *“…So we try and do an induction, give people the chance to observe and if they can’t do that we will do an induction but maybe that might be something that would be good to keep so they get to know our faces and they get to know that they can come to us still although it is remote….” (P2)*

## DISCUSSION

To our knowledge this is the first study to have explored funding committee practices through observations, complemented with follow up interviews and an online survey with committee members, and interviews with staff who facilitated and organised these meetings. The study also applied a methodological approach that specifically focuses on online social interactions, which offered a unique and in-depth understanding about the social processes of virtual funding committees given their central role in the funding process. Examining the decision-making practices of one funding organisation across several research programmes and across multiple committee meetings over a period of one year has therefore generated new insights around funding committee practices that previous studies have not been able to explore or investigate due to gaining access or sensitivities around the funding allocation processes.^(2–6)^

The findings highlighted the complexities of preparing and running funding committee meetings and also how the meetings, when conducted virtually, introduce new challenges and benefits than those conducted in a different setting (e.g., face-to-face).^(6, 11)^ The study found that several parameters are not transferable from a face-to-face to a virtual setting, such as timings, location, equipment, and physical attributes. All of which can have wider implications for funding committee members and funding organisations preparing and structuring the committee meetings. Virtual meetings require different functional considerations, such as how to manage conflicts of interest when someone cannot simply leave the room or when someone has a poor internet connection. The findings highlight the level of planning and preparation required by staff to mitigate against these issues along with the repercussions of being reliant on technology, meant that more concentration was required from all staff and committee members, and often resulted in an increased level of fatigue. In addition, scheduled breaks and changes to the standard structure of committee meetings resulted in less free time, further exacerbating fatigue in the virtual experience.

Adjusting to using virtual platforms during the pandemic has shown that over time, committee members and staff have become more accepting of new ways of working, as what was seen as ‘daunting’ in the latter part of 2020, was far less of an issue one year on. Whilst it was evident that running committee meetings virtually had its benefits in terms of work-life balance, travel, and environmental sustainability, it was suggested that this sometimes came at a price. For many, there was a ‘trade off’ with not having the opportunity to socially interact or network whilst attending the funding committee meeting, as well as increased fatigue. The themes highlight how important levels of engagement and social interaction are, especially for new members of the committee and during committee discussions. These findings are in line with previous literature, suggesting that an unequal focus or limited engagement from funding committee members can lead to or limit funding decision discussions.^(6, 11, 25)^ Nevertheless, observations of committees and feedback from committee members agreed that despite discussions being more focused, the decision-making process was largely the same in the virtual meeting.

### Future considerations and recommendations

The role and function of committee meetings, whether they are virtual or face-to-face does not change, and both have benefits and challenges. It is important to note that some of the challenges reported about virtual committees were also relevant to face-to-face meetings. For example, the potential for unconscious bias was not something unique to virtual meetings and is reported in the literature.^(25, 26)^ What was evident from the observations was how the virtual committees evolved over time and adapted their approach to accommodate committee members but also to make the process easier to manage (**see supporting materials S1 Table: Considerations and recommendations for future virtual funding committees).**

The organisational structure of funding committee meetings breaks and timings of the applications need careful consideration, with potential flexibility or options to change the agenda order. This is especially relevant to committees who have members from different time zones. It is also important that processes are in place to minimise potential biases and ensure no power imbalances between different committee members or towards certain applications. As demonstrated by one of the NIHR funding committees observed, one possible consideration that may help with this is for all members of the committee to view an unconscious bias video prior to reviewing applications as part of the preparation process of the funding committee meeting. More broadly, there is also an opportunity for funding organisations to consider a more inclusive and diverse funding committee membership that takes account of differences in time zones, disabilities, part-time work, or those with other responsibilities.

The process and formality of running funding committee meetings is also imperative to ensure inclusive contributions and engagement for all members of the committee. Encouraging members to have their cameras on is one consideration, although there are challenges associated to this, especially where internet connections are unreliable. There is a need to appreciate how new members interact and engage with existing funding committees to encourage participation and contribution to the discussions. Such opportunities could consider face-to-face development days or a mixed approach to how the funding committees are held (e.g., mixture of face-to-face and virtual meetings).

Finally, future requirements for training and additional guidance to support existing and new committee members and the chair are important considerations. Chairing virtual meetings is different and requires a different set of skills. At times it was challenging for the chairs, who found it difficult due to having limited social cues from the committee to aid discussion, multiple technologies/screens to manage and because the duration of the meetings meant that they had to interact virtually for extended periods of time. This often led to increased fatigue, particularly during the first round of committee meetings. Several options could be considered to lessen the virtual fatigue, including guidance to support committee members and chairs, extended breaks and/or shorter meetings. It is also important to consider how new members of the funding committee could be integrated into existing committees, when the format is predominately virtual. Such considerations could be face-to-face development days or virtual social meetings. In addition, allowing more junior members to join as observers or trainees on a funding committee may encourage diversification of funding committees as a form of training. Thus, virtual funding committees not only have additional training considerations but also the offer the opportunity to be a form of training which in turn may facilitate the diversity of committee membership and increase the transparency around funding committee practices.

### Strengths and limitations

The main strength of the study was the inclusion of nine NIHR committee meetings across several research funding programmes. As we included committee meetings that took place over a one-year period, it meant that committee members and staff had experienced more than one virtual committee meeting. This enabled us to see how the views, opinions and expectations of committee members changed over time. Capturing these experiences had important implications for the findings and how experiences can vary across different funding programmes. As the study included interviews and an online survey, we were able to follow up and support our non-participatory observational claims, which can often be seen as a limitation of netnographic studies. It is also important to note that due to the complexity, structure and formality of funding committee meetings, some areas considered important ran through more than one theme. By using a methodological approach that was based around online social interactions, it was possible to gain valuable insights into the recommendations of research funding allocation, without influencing the views, opinions, or expectations of the committees or staff.

Ethical considerations and recommendations of netnographic studies are important to ensure information about users’ identification is kept confidential, which is frequently reported in the literature as a weakness of this type of study.^(15, 27)^ To overcome this, the study sought ethical approval and had a high cloaking level to avoid identification of the survey respondents, interviewees and members of the committee.

A limitation to the study was that it was based on observations and experiences of funding committee meetings held in the early part of the COVID-19 pandemic (2020-2021) and therefore did not include any comparable data with face-to-face committee meetings. Early insights to the findings of the study and based on committees own experience, some changes to the funding committee process may have already been implemented due to developments and initiatives taking place simultaneously with the current study. Another limitation was around potential researcher bias during analysis and interpretation of the observations, interviews, and survey responses. Four researchers involved in the analysis, each bringing their own experiences and knowledge on funding committee practices, could have produced bias on overall expectations and interpretation of findings. However, actively encouraging to keep an immersive journal throughout data collection and analysis, and enabling regular reflective discussions on reactions, feelings, and observations, helped to minimise bias and maintain a level of autonomy.

## CONCLUSION

Although there are several areas for consideration for continued virtual funding committee meetings, such as inducting new members and maintaining inclusivity for all committee members, the study found that conducting funding committee meetings virtually was feasible and funding decisions continued to be fair and transparent.

Given that there is no current evidence or use of observations to understand the social processes and functions of funding committee meetings, this study has shown its value and critical contribution to building an evidence-informed approach. By applying a netnography methodology to observe, understand and capture the views of virtual funding committees, it was possible to gain insight to these committees attended by the respondents.

Although there is acceptance and a place for virtual committee meetings from committee members and staff, it is important to remember that this is not the view of all members. Whilst virtual funding committees have many benefits and opportunities such as the potential to enable work-life balance, inclusivity for members, reduce costs, and be more environmentally sustainable, more evidence is needed to evaluate the longer-term sustainability of virtual committee meetings in the allocation and decision-making of funded research.

## Data Availability

In line with our ethical approval, the interview, survey, and observational data cannot be shared publicly to maintain confidentiality of our participants. The interview, survey and observational guides are provided in the Supporting Information files. Additional quotes under each theme are available from the Insight team, School of Healthcare Enterprise and Innovation, University of Southampton (contact via or corresponding author) for researchers who meet the criteria for access to the data.

## Supporting information

**S1 Appendix**. Observation guide

**S2 Appendix**. Interview guide for committee members

**S3 Appendix**. Interview guide for NIHR staff

**S4 Appendix**. Survey questions

**S1 Table**. Future considerations and recommendations for funding committee meetings

## Acknowledgements

We would like to thank all participants that took part in the study and for those NIHR staff who contributed to the early design of the study. We would also like to thank the NIHR Research on Research team for reviewing early draft versions of the article.

## Abbreviations

AI: Artificial Intelligence
COI: Conflict of Interest
COVID-19: Coronavirus Disease 2019
DCM: Designated Committee Member/s
EME: Efficacy and Mechanism Evaluation
ES: Evidence Synthesis
GHR: Global Health Research
HSCDS: Health and Social Care Delivery Service
HSDR: Health Service and Delivery Research
HTA: Health Technology Assessment
NIH: National Institutes of Health
NIHR: National Institute for Health and Care Research
NIHRCC: National Institute for Health and Care Research Coordinating Centre
P1: Survey written quotations
P2: Interview verbal quotations
P3: Observation verbal quotations
PGfAR: Programme Grants for Applied Research
PHR: Public Health Research
PIS: Participant Information Sheet
PPIE: Patient and Public Involvement and Engagement
RfPB: Research for Patient Benefit

## Author contributions

**Conceptualization:** Amanda Blatch-Jones, Katie Meadmore

**Data curation:** Amanda Blatch-Jones, Katie Meadmore

**Formal analysis:** Amanda Blatch-Jones, Katie Meadmore, Emmanuel Asante, Cherish Boxall

**Investigation:** Amanda Blatch-Jones, Katie Meadmore, Emmanuel Asante, Cherish Boxall

**Methodology:** Amanda Blatch-Jones

**Project administration:** Amanda Blatch-Jones

**Supervision:** Amanda Blatch-Jones

**Validation:** Amanda Blatch-Jones, Katie Meadmore

**Visualization:** Amanda Blatch-Jones

**Writing – original draft:** Amanda Blatch-Jones, Katie Meadmore

**Writing – review and editing:** Amanda Blatch-Jones, Katie Meadmore, Emmanuel Asante, Cherish Boxall

